# Inequities in childhood anaemia in Mozambique: results from multilevel Bayesian analysis of 2018 National Malaria Indicator Survey

**DOI:** 10.1101/2021.03.24.21252471

**Authors:** Nazeem Muhajarine, Daniel Adedayo Adeyinka, Mbate Matandalasse, Sergio Chicumbe

## Abstract

**Introduction:** Childhood anaemia is a common public health problem worldwide. The geographical patterns and underlying factors of childhood anaemia have been understudied in Mozambique. The objectives of this study were to identify the child-, maternal-, household-, and community-level determinants of anaemia among children aged 6-59 months, and the contribution of these factors to the variation in childhood anaemia at the community level in Mozambique.

**Methods:** This is a cross-sectional study that utilized data of a weighted population of 4,141 children aged 6-59 months delivered by women between 15-49 years of age, from the 2018 Mozambique Malaria Indicator Survey. Multilevel Bayesian linear regressions identified key determinants of childhood anaemia. Spatial analysis was used to determine geographic variation of anaemia at the community level and areas with higher risks.

**Results:** The overall national prevalence of childhood anaemia was 78-80.3%. There was provincial variation with Cabo Delgado province (86.2%) having highest prevalence, and Maputo province (70.2%) the lowest. Children with excess risk were mostly found in communities that had proximity to provincial borders: Niassa-Cabo Delgado-Nampula tri-provincial border, Gaza-Inhambane border, Zambezia-Nampula border, and provinces of Manica and Inhambane. Children with anaemia tended to be younger, males, and at risk of having malaria because they were not sleeping under mosquito nets. In addition, children from poor families and those living in female-headed households were prone to anaemia.

**Conclusion:** This study provides evidence that anaemia among children aged 6-59 months is a severe public health threat across the provinces in Mozambique. It also identifies inequity in childhood anaemia—worse among communities living close to the provincial borders. We recommend interventions that would generate income for households, increase community-support for households headed by women, improve malaria control, build capacity of healthcare workers to manage severely anaemic children and health education for mothers. More importantly, there is need to foster collaborations between communities, districts and provinces to strengthen maternal and child health programmes for the severely affected areas.

**What is already known?:**

- Nearly two billion people are anaemic, affecting mostly poor women and children. Anaemia, a co-morbidity with other major health conditions, frequently is less prioritized.
- Sustainable Development Goals 2 and 3, formulated to tackle hunger/food insecurity and attain optimal health/wellbeing, respectively, currently have no specific target for monitoring global progress for anaemia among children.

**What are the new findings?:**

- Twenty-four percent of children (6-59 months) had anaemia classified as mild, 50% moderate and 7% severe.
- Childhood anaemia showed spatial variation across the communities—especially in the provincial border regions--and provinces in Mozambique; they were younger, males, at risk of having malaria, from poor families and lived in female-headed household.

**What do the new findings imply?:**

- Anaemia among children could be effectively reduced through malaria prevention, e.g. bed netting.
- This report of anaemia at community and district level provides baseline data and can guide targeted implementation of the 2025 Mozambique National Development Plan.
- Interventions needed that generate income for households, increase community-support for households headed by women, improve malaria control, build capacity of healthcare workers to manage severely anaemic children and health education for mothers.

## Introduction

Anaemia—a condition typically defined as shortage of red blood cells or haemoglobin of less than the 5^th^ percentile for the individual’s age (i.e., ≤5g/dl or 11.5%), is a common public health problem worldwide.^(1)^ Nearly two billion people are anaemic, affecting mostly poor women and children— corresponding to one-third of the world’s population.^(2)^ Despite the huge physiological and socio-economic impacts of anaemia in the recent decades, it has attracted less attention by policymakers.^(3)^ Moraleda et al.,^(3)^ argued that the long neglect of anaemia could be partly due to its co-morbidity with other major health conditions, which are frequently more prioritized. Moreover, although the Sustainable Development Goals 2 and 3 were formulated to tackle hunger/food insecurity, and maintain optimal health/wellbeing for people, respectively,^(4,5)^ there is currently no clear target for monitoring global progress for anaemia among children.

Recent findings from a systematic analysis of 328 diseases in 195 countries highlighted that iron-deficiency anaemia is among the first five leading causes of years lived with disability (DALY).^(6)^ The negative societal impacts of anaemia are more pronounced in the low- and middle-income countries (LMICS), where it causes premature morbidity and mortality of pregnant women and under-five children, resulting in significant human capital and economic losses.^(6)^ As previously reported in literature, anaemia impairs cognitive functioning during the early childhood developmental stage,^(7)^ and those children affected often suffer from other complications of iron and vitamin B12 deficiencies.^(8–10)^ The main long-term consequences are poor psychomotor skills, academic performances and susceptibility to infections.^(8–10)^ The need to urgently tackle global food insecurity is therefore a top priority as studies have acknowledged that poor nutrition is the most common cause of anaemia worldwide.^(8–10)^ Other important proximal contributors to childhood anaemia in sub-Saharan Africa are malaria, haemoglobinopathies, HIV, tuberculosis, and intestinal worm infestations.^(8,11–13)^

The global prevalence of anaemia among under-five children has remained at 41.7% between 2012 and 2016.^(14)^ According to World Health Organization (WHO), Mozambique is one of the sub-Saharan African countries with high prevalence of childhood anaemia—60% in 2016, ranking 26^th^ in league tables.^(14)^ The government of Mozambique is currently making efforts to address nutritional anaemia among women and the children through programs specifically targeted to these populations—such as mass deworming, iron supplementation and malaria chemoprophylaxis at antenatal care—as well as by providing emergency food assistance, food vouchers and nutrition counseling to populations in need.^(15)^ However, findings from the 2011 Mozambique Demographic and Health Survey (MDHS) showed that food insecurity persisted, 63% of under-five children showing signs of moderate to severe chronic malnutrition.^(16)^ Malnutrition is also an underlying cause of about 26% of under-five deaths in Mozambique.^(17)^

Some studies in sub-Saharan Africa have recognized the individual- and community-level factors associated with childhood anaemia,^(18,19)^ however, the key determinants of childhood anaemia and its variation across the provinces of Mozambique are less understood. Most of the previous studies are hospital-based and extensively focused on medical causes of childhood anaemia, with less attention to its social determinants,^(3)^ or utilized data from multiple countries,^(19,20)^ making evidence needed for action difficult. In one of the few nationwide studies in Mozambique, Mabunda et al.,^(21)^ concluded that there was no significant difference in the provincial mean haemoglobin concentration among children under 10 years old. Adeyemi et al.,^(22)^, however, observed a significant spatial pattern of childhood anaemia—worse in central Mozambique in 2011. Given the scarce evidence needed to improve childhood anaemia in the communities and districts, this study is aimed to broaden the extant knowledge on childhood anaemia in Mozambique by analysing the newly released 2018 Malaria Indicator Survey (MIS). The objectives of this study were to identify the child-, maternal-, household-, and community-level determinants of anaemia among children aged 6-59 months, and the contribution of factors to the variation in childhood anaemia at the community level in Mozambique.

## Methods

### Data source

This is a cross-sectional study that utilized child recode datafile and the Global Positioning System (GPS) dataset of the 2018 Mozambique Malaria Indicator Survey.^(23)^ The survey was conducted between April 2018 and June 2018 by the Moçambique Instituto Nacional de Saúde, in collaboration with ICF International Calverton, Maryland, USA, to provide national and subnational estimates of anaemia and malaria indicators for policy and programmatic purposes. The details of the methodology used for the survey has been published elsewhere.^(24)^ With a stratified two-stage sampling design, face-to-face standardized questionnaire interviews were conducted among women aged 15-49 years. Using probability proportional to size, the first stage of sampling involved selection of 224 clusters or enumeration areas (otherwise known as the primary sampling units (PSU)) from the 2007 General Population and Housing Census.^(24)^ Out of the 224 clusters, 58.9% were in rural areas, while the rest were in urban areas. The second stage involved systematic sampling of 6,279 households. Each household was randomly selected from the household listing, with an average of 28 households per cluster. For this study, the clusters are referred to as the “communities” because they are believed to comprise homogenous or kinship populations. Of the 6,279 households, the response rate was 99%, while out of the 6,290 eligible women identified, 6,184 could be interviewed (response rate of 98.3%). With the consent of the parents/caregivers, blood samples were collected in a microcuvette from the heel (of children aged 6-11 months) or fingers (of the children aged 12-59 months) for haemoglobin concentration estimation. After discarding the first drop of blood to avoid possible contamination, haemoglobin concentration was estimated with the second drop, using an automatic hematology analyzer (HemoCue® 201+). For this study, a subpopulation of 3,843 (weighted: 4,141) singletons aged 6-59 months who were alive at the commencement of the survey were analyzed.

### Ethical considerations

This is a secondary analysis of data available online(23) where the datasets are de-identified of the respondents’ personal information, hence no ethical approval was required for this specific study. However, prior to the commencement of the primary survey, ethical clearances were obtained by the Demographic and Health Survey (DHS) team from National Committee for Bioethics in Health of Mozambique (Comité Nacional de Bioética para Saúde, CNBS). Also, written informed consent was obtained from all mothers during the field work. Following registration, administrative access to the dataset was granted by the DHS team, United States. The survey data files were provided at no cost for academic research.(25)

### Variables

#### Outcome variable

The outcome variable was child’s anaemic status, measured as altitude-adjusted hemoglobin (Hb) concentration (in g/L), and the severity of anaemia was categorized based on pre-defined threshold values: normal (≥110 g/L), mild (100-109 g/L), moderate (70-99 g/L) and severe (<70 g/L).^(26)^

#### Exposure variables

The selection of exposure variables was guided by the socio-ecological model,^(27)^ and on the evidence that the proposed variables are important covariables to be included in relation to the outcome.

The socio-ecological model proposes that health outcomes are influenced by factors that operate at different levels i.e., micro-level (individual), meso-level (household), and macro-level (community/district/province). The independent variables comprise of factors that affect the outcome at different levels, e.g. child-level factors—age (in months), sex (male and female), birth order (surviving eldest child and non-first-born children), mosquito net usage—as a proxy for risk of malaria illness—(no net, only untreated nets and only treated nets), preceding birth interval (<24 months and ≥24 months) and succeeding birth interval (<24 months and ≥24 months); maternal-level factors—age in years (≤19, 20-34 and ≥35), highest maternal educational attainment (no education, primary, secondary and post-secondary), parity (≤2, 3-5 and >5) and adequacy of antenatal care (ANC) visits based on WHO guidelines^(28)^—none, inadequate (1-7 visits) and adequate (≥8 visits); household-level variables—sex of household head (male and female), wealth index—proxy for food (in)security. The wealth index is a composite measure of household’s assets and amenities (e.g., ownership of television, bicycle, car, livestock etc.), which was calculated using principal components analysis. The standardized scores were divided into quintiles. We recoded MIS wealth index into low (i.e., poor and poorest), middle, and high (i.e., rich and richest) socioeconomic status. Based on the WHO/UNICEF Joint Monitoring Programme for Water Supply, Sanitation and Hygiene (JMP) classification,^(29)^ household sanitation, source of drinking water, and source of cooking fuel (proxy for indoor pollution) were recoded as improved and unimproved. The community-level variables included were social infrastructural development (measured by proportion of households with electricity in the community), place of residence (urban and rural) and province.

### Statistical analysis

The unit of statistical analysis was the individual child. Following the preliminary data quality checks, there was no multicollinearity among the independent variables (mean variance inflation factor (VIF)=1.29). For the descriptive analyses, we used the complex survey analysis commands in Stata™ software version 16.1^(30)^ to account for the cluster sampling design and applied the sampling weights provided in the datafile.

Considering the nested nature of the data, we used multilevel Bayesian linear regression with four-level models. Practically, modelling haemoglobin concentration as a continuous outcome variable prevented misclassification and allowed for retention of maximum information in the linear regression, which could have been lost with categorization.^(31)^ In order to maximize simulation efficiency and reduce the long computational time often required to achieve convergence for multilevel Bayesian models, the outcome variable (haemoglobin concentration) was log-transformed.^(30)^ Also, two Monte Carlo Markov Chains (MCMC) were simulated with a hybrid algorithm of random-walk Metropolis–Hastings and Gibbs sampling to improve the precision of parameter estimates.^(30)^ The prior specifications for the regression parameters were set to follow a non-informative normal distribution—zero mean and a variance of 10,000, because there was no prior knowledge of the directions and effect sizes of the independent variables. We fitted 25,000 iterations per chain (including a burn-in period of 5,000 iterations per chain), hence generating a MCMC sample size of 40,000 iterations with an acceptable Monte Carlo errors (<5% of the posterior standard deviation). Convergence of MCMC samples is crucial in making reliable inferences from Bayesian analysis, hence, monitoring was done with a strict convergence rule of Gelman-Rubin statistic Rc <1.1.^(30)^ In addition, the Markov chains were checked by visually inspecting the histograms, trace, autocorrelation, and kernel density plots (Supplementary Figures S1-S5).

#### Model building

We fitted five models. Model 0 was an empty/null model which did not include any covariate, but specified three-random intercepts attributable to mothers, households and communities. Model 1 added child-level variables to Model 0. Using parsimonious approach, Model 2 retained variables from Model 1 and added maternal-level variables. Model 3 comprised variables retained from Model 2 and added household-level factors. Model 4 (full model) retained variables in Model 3 and added community-level variables. The goodness-of-fit of final main effect model was evaluated with an average Bayesian Deviance Information Criterion (DIC), using Laplace-Metropolis approximation.^(30)^ All Bayesian models were fitted in the Stata 16.1 software.^(30)^

### Measures of associations (fixed effects)

The measures of association were presented as posterior exponential ⍰ coefficients and 95% Bayesian Credible Intervals (95%CIs). The level of statistical significance was determined by the non-inclusion of one in the 95%CIs.

### Measures of variations (random effects)

To evaluate the extent of variation of childhood anaemia among the women, households and communities, intraclass correlation coefficient (ICC), median odds ratio (MOR) and proportional change in variance (PCV) were calculated.

The ICC at the maternal, household and community levels were estimated as shown in equations i, ii and iii, respectively.^(31)^

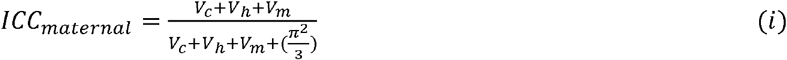

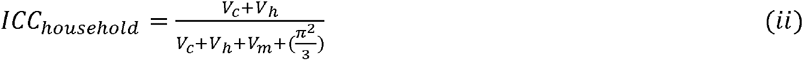

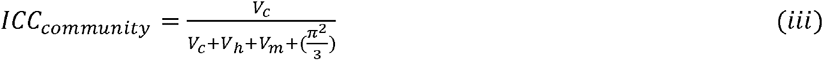

*Where V*_*m*_, *V*_*h*_ *and V*_*c*_ *are the variances of childhood anaemia at the maternal, household and community levels, respectively*.

The MOR was estimated to measure the unexplained heterogeneity of childhood anaemia at a specified level—maternal, household and community. Using equation (iv), MOR measured the probability of having anaemia when a child is randomly picked from a setting of lowest risk to highest risk.^(32)^

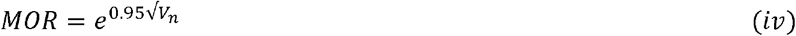

*Where V*_*n*_ *is the variance at specified level, i*.*e*., *maternal, household and community*.

The PCV estimated the total variance attributable to the independent variables for each level of the multilevel model.

#### Spatial analyses

##### Rates-calculated maps

Initially, we generated unadjusted provincial prevalence maps (excess risk and spatial empirical Bayesian smoothed maps) in GeoDa v.1.14.^(33)^ The smoothed provincial map was based on geometric centroids and spatial (arc) distance of 396.16km in a geographic coordinate map (WGS84: EPGS4326). The empirical Bayesian smoothed prevalence map accounted for the intrinsic variance instability of small area disease risks associated with complex survey data, by minimizing the standard errors.^(34)^ The smoothing is important to avoid spurious representation of the spatial patterns of the risk of childhood anemia for Mozambique. The following equations were used to account for spatial heterogeneity and spatial dependence between provinces.^(34)^

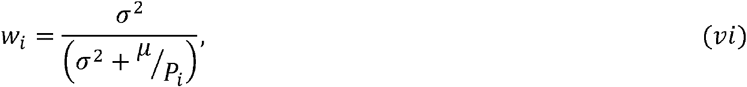

Where 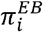 is the empirical Bayesian estimate for the risk in province i, θ is the prior estimate, r is the weighted average of crude rate.

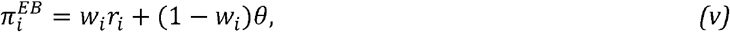

Where P_i_ is the population at risk in province i, and μ and ⍰^2^ are the mean and variance of the prior distributions.

##### Identification of hot spots

We linked the weighted cases of anaemia and weighted population-at-risk to each community (vector points) in the 2018 Mozambique MIS geographic shapefile and determined the presence of geographic clustering (positive spatial autocorrelation) with global Moran’s I index and local indicator spatial autocorrelation (LISA) maps at a Monte Carlo Randomization (MCR) of 999 permutations and p-value <0.05.

For comparison purpose, the crude and adjusted posterior means of the provincial-level random effects from the multilevel Bayesian linear regression were mapped. In order to predict the mean haemoglobin concentration for other communities that were not captured in the 2018 MIS, we used spatial interpolation with inverse weighing distancing in QGIS v.2.8.9 software.^(35)^ With isopleth maps, it is easier to identify the districts with different posterior means of haemoglobin concentration.

## Results

### Sample characteristics

Table 1 summarizes the sample characteristics. The mean age (standard deviation (SD)) of the children in the study was 31.2 (±17.6) months, with a male to female ratio of 1.06:1. Sixty-seven percent of mothers of children in the study were between 20-34 years of age. On average, mothers had 4 children. Almost half (49%) of the children lived in poor households (ranging from 3.01% in Maputo city to 20.4% in Nampula)—similar to the 48% of children living in absolute poverty reported in the 2011 UNICEF report on child poverty and disparities in Mozambique.^(36)^ Most children’s mothers had not finished or had only primary school education (79.46%); 97% households used polluting fuels for cooking, 73% resided in rural communities, and 72% lived in households that were headed by men.

**Table 1:**
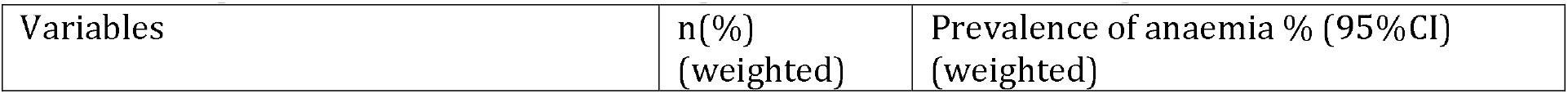
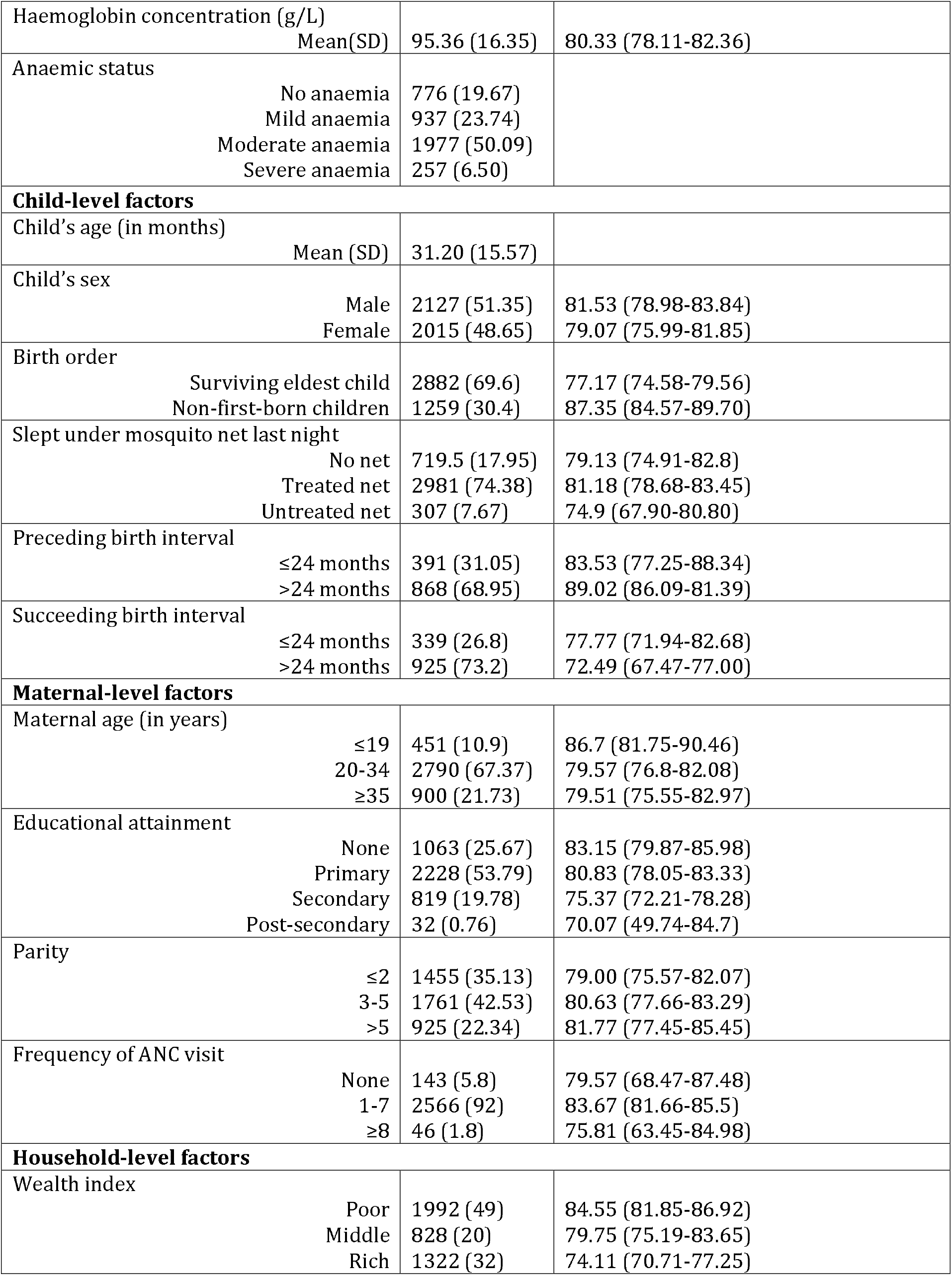
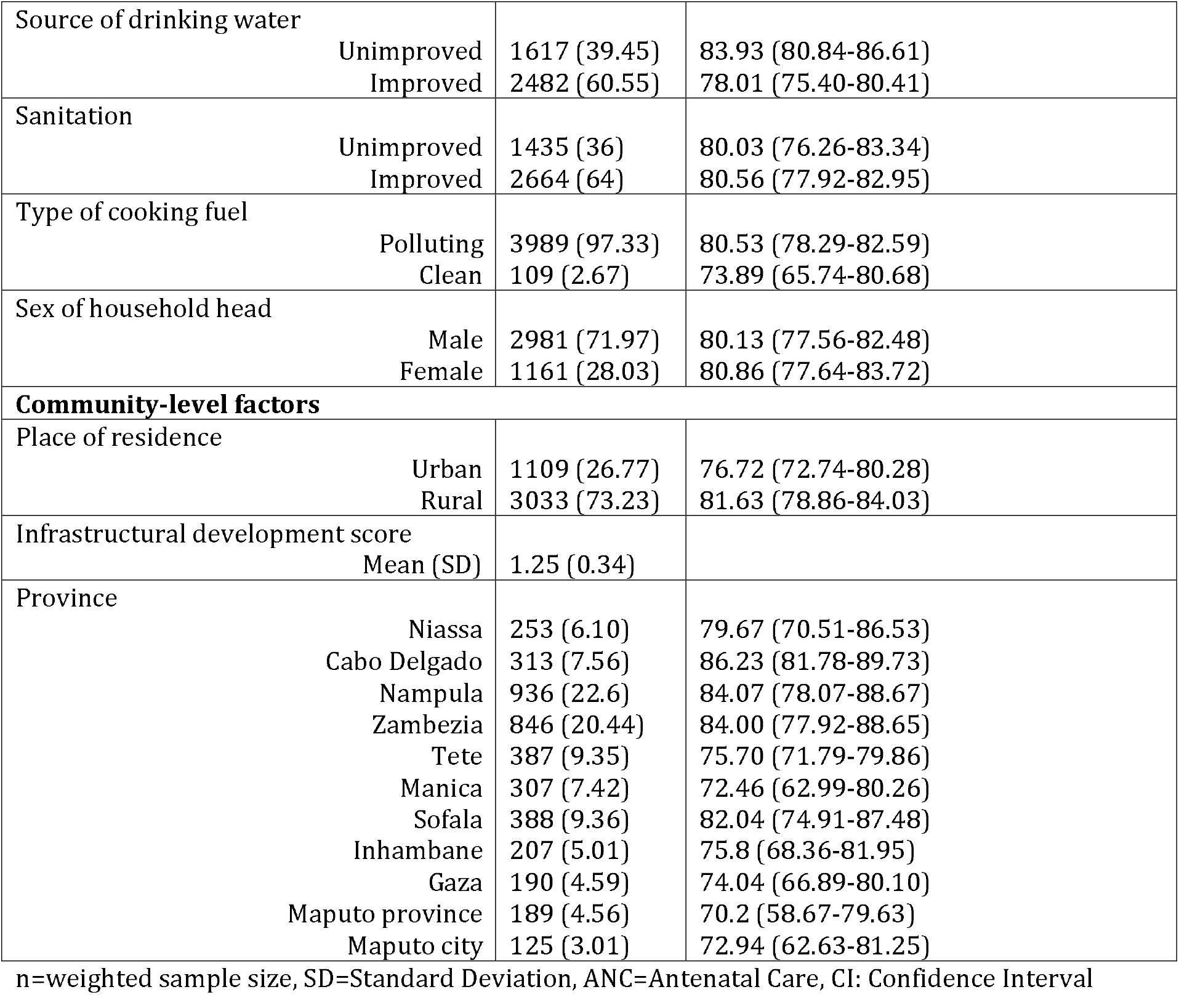
Sample characteristics of children aged 6-59 months, Mozambique MIS 2018

| Variables | n(%)<br>(weighted) | Prevalence of anaemia % (95%CI)<br>(weighted) |
| --- | --- | --- |
| Haemoglobin concentration (g/L) |  |  |
| Mean(SD) | 95.36 (16.35) | 80.33 (78.11-82.36) |
| Anaemic status |  |  |
| No anaemia | 776 (19.67) |  |
| Mild anaemia | 937 (23.74) |  |
| Moderate anaemia | 1977 (50.09) |  |
| Severe anaemia | 257 (6.50) |  |
| **Child-level factors** |  |  |
| Child's age (in months) |  |  |
| Mean (SD) | 31.20 (15.57) |  |
| Child's sex |  |  |
| Male | 2127 (51.35) | 81.53 (78.98-83.84) |
| Female | 2015 (48.65) | 79.07 (75.99-81.85) |
| Birth order |  |  |
| Surviving eldest child | 2882 (69.6) | 77.17 (74.58-79.56) |
| Non-first-born children | 1259 (30.4) | 87.35 (84.57-89.70) |
| Slept under mosquito net last night |  |  |
| No net | 719.5 (17.95) | 79.13 (74.91-82.8) |
| Treated net | 2981 (74.38) | 81.18 (78.68-83.45) |
| Untreated net | 307 (7.67) | 74.9 (67.90-80.80) |
| Preceding birth interval |  |  |
| ≤24 months | 391 (31.05) | 83.53 (77.25-88.34) |
| >24 months | 868 (68.95) | 89.02 (86.09-91.39) |
| Succeeding birth interval |  |  |
| ≤24 months | 339 (26.8) | 77.77 (71.94-82.68) |
| >24 months | 925 (73.2) | 72.49 (67.47-77.00) |
| **Maternal-level factors** |  |  |
| Maternal age (in years) |  |  |
| ≤19 | 451 (10.9) | 86.7 (81.75-90.46) |
| 20-34 | 2790 (67.37) | 79.57 (76.8-82.08) |
| ≥35 | 900 (21.73) | 79.51 (75.55-82.97) |
| Educational attainment |  |  |
| None | 1063 (25.67) | 83.15 (79.87-85.98) |
| Primary | 2228 (53.79) | 80.83 (78.05-83.33) |
| Secondary | 819 (19.78) | 75.37 (72.21-78.28) |
| Post-secondary | 32 (0.76) | 70.07 (49.74-84.7) |
| Parity |  |  |
| ≤2 | 1455 (35.13) | 79.00 (75.57-82.07) |
| 3-5 | 1761 (42.53) | 80.63 (77.66-83.29) |
| >5 | 925 (22.34) | 81.77 (77.45-85.45) |
| Frequency of ANC visit |  |  |
| None | 143 (5.8) | 79.57 (68.47-87.48) |
| 1-7 | 2566 (92) | 83.67 (81.66-85.5) |
| ≥8 | 46 (1.8) | 75.81 (63.45-84.98) |
| **Household-level factors** |  |  |
| Wealth index |  |  |
| Poor | 1992 (49) | 84.55 (81.85-86.92) |
| Middle | 828 (20) | 79.75 (75.19-83.65) |
| Rich | 1322 (32) | 74.11 (70.71-77.25) |

|  |  |  |  |
| --- | --- | --- | --- |
| Source of drinking water | Unimproved | 1617 (39.45) | 83.93 (80.84-86.61) |
|  | Improved | 2482 (60.55) | 78.01 (75.40-80.41) |
| Sanitation | Unimproved | 1435 (36) | 80.03 (76.26-83.34) |
|  | Improved | 2664 (64) | 80.56 (77.92-82.95) |
| Type of cooking fuel | Polluting | 3989 (97.33) | 80.53 (78.29-82.59) |
|  | Clean | 109 (2.67) | 73.89 (65.74-80.68) |
| Sex of household head | Male | 2981 (71.97) | 80.13 (77.56-82.48) |
|  | Female | 1161 (28.03) | 80.86 (77.64-83.72) |
| **Community-level factors** |  |  |  |
| Place of residence | Urban | 1109 (26.77) | 76.72 (72.74-80.28) |
|  | Rural | 3033 (73.23) | 81.63 (78.86-84.03) |
| Infrastructural development score |  |  |  |
| Mean (SD) |  | 1.25 (0.34) |  |
| Province | Niassa | 253 (6.10) | 79.67 (70.51-86.53) |
|  | Cabo Delgado | 313 (7.56) | 86.23 (81.78-89.73) |
|  | Nampula | 936 (22.6) | 84.07 (78.07-88.67) |
|  | Zambezia | 846 (20.44) | 84.00 (77.92-88.65) |
|  | Tete | 387 (9.35) | 75.70 (71.79-79.86) |
|  | Manica | 307 (7.42) | 72.46 (62.99-80.26) |
|  | Sofala | 388 (9.36) | 82.04 (74.91-87.48) |
|  | Inhambane | 207 (5.01) | 75.8 (68.36-81.95) |
|  | Gaza | 190 (4.59) | 74.04 (66.89-80.10) |
|  | Maputo province | 189 (4.56) | 70.2 (58.67-79.63) |
|  | Maputo city | 125 (3.01) | 72.94 (62.63-81.25) |

n=weighted sample size, SD=Standard Deviation, ANC=Antenatal Care, CI: Confidence Interval

### Prevalence of anemia

Overall, four out of five children had anaemia, translating to a national prevalence of 80.33% (95%CI: 78.11-82.36%) (Table 1). The mean (±SD) haemoglobin concentration was 95.4 (±16.4) g/L. Twenty-four percent of children had anaemia classified as mild, 50% moderate and 7% severe. The prevalence of anaemia was clearly high in males (81.5%), those born late in the birth order (87.4%), born to mothers who had children closer together (less than two-year gap)—77.8%, delivered by teenage mothers (86.7%), living in houses with unimproved sources of drinking water (83.9%), and residing in rural communities (81.6%). The prevalence of anaemia was higher in children with mothers without formal education (83.2%), delivered by women who had more than five births (81.8%), and those from poor households (84.6%). Across the provinces, Cabo Delgado had the highest prevalence of childhood anaemia (86.2%) and lowest in Maputo province (70.2%).

### Factors associated with childhood anaemia

Figures 1a-d present the results of the fixed-effect estimates from the multilevel Bayesian linear regression for child-level factors (Model 1), child and maternal-level factors (Model 2), child, maternal and household-level factors (Model 3) and child, maternal, household and community-level factors (Model 4). From the final multivariate model, the haemoglobin concentration was 2.2% higher for female children than for male children (posterior exp(⍰): 1.022, 95%CI: 1.010-1.033) (Figure 1d). For a ten-unit increase in child’s age in months, there was a 3% increase in haemoglobin concentration (posterior exp(⍰): 1.003, 95%CI: 1.003-1.003). For a one-unit increase in the number of children sleeping under mosquito net, there was a 2.8% increase in the mean haemoglobin concentration, compared to those not sleeping under mosquito net (posterior exp(⍰): 1.028, 95%CI: 1.002-1.055). A dose-response relationship was observed for household wealth and haemoglobin concentration. The percentage increment in mean haemoglobin concentration was 2.8% in children of households with middle-wealth index (posterior exp(⍰): 1.028, 95%CI: 1.010-1.029), 5.4% in children of households with rich-wealth index (posterior exp(⍰): 1.054, 95%CI; 1.029-1.079), compared to those from poor households. Children living in households headed by women, compared to those living in households with male-heads, had 1.3% decrease in mean haemoglobin concentration (posterior exp(⍰): 0.987, 95%CI: 0.973-1.000). Compared to children living in Maputo province, those from Cabo Delgado province had the largest reduction (8.2%) in mean haemoglobin concentration (posterior exp(⍰): 0.918, 95%CI: 0.875-0.964), followed by 8.1% drop in Sofala province, posterior exp(⍰): 0.919, 95%CI: 0.878-0.961. There was 6.8% decrease in mean haemoglobin concentration for Nampula province, posterior exp(⍰): 0.932, 95%CI: 0.890-0.975; 6.7% decline in Niassa, posterior exp(⍰): 0.933, 95%CI: 0.890-0.977, and 6.5% decrease in Zambezia province, posterior exp(⍰): 0.935, 95%CI: 0.892-0.979).

**Figure 1:**
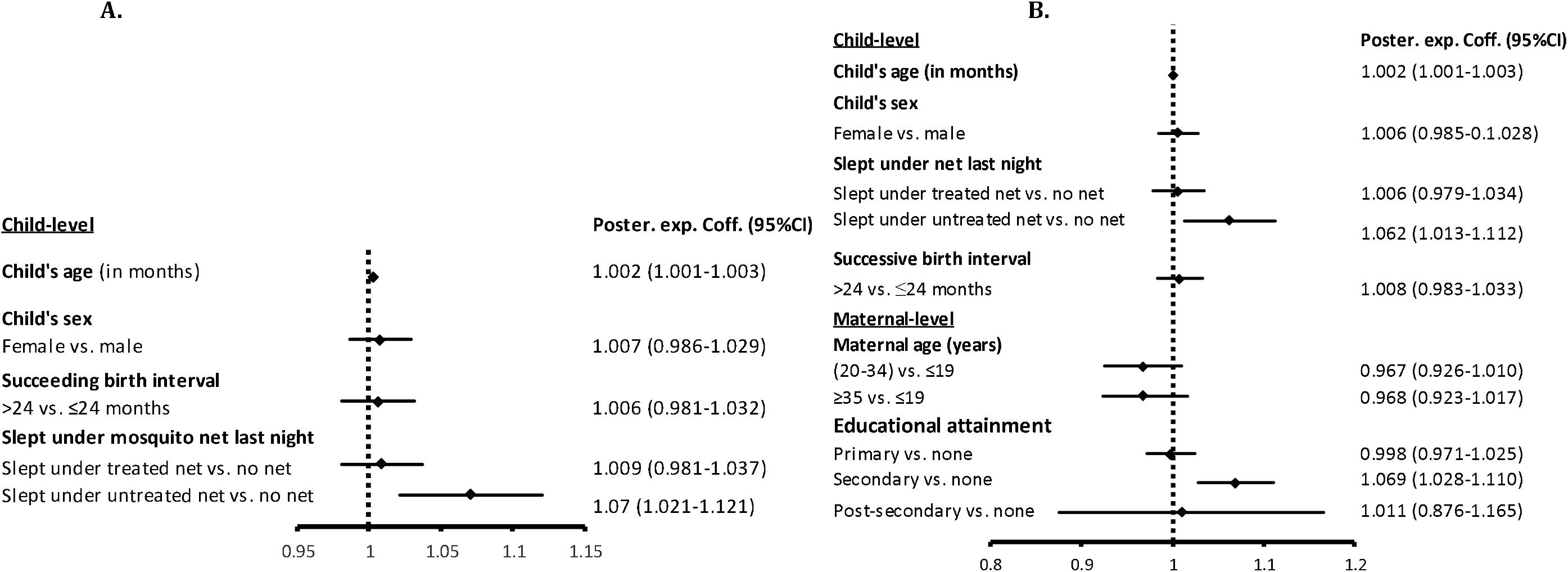
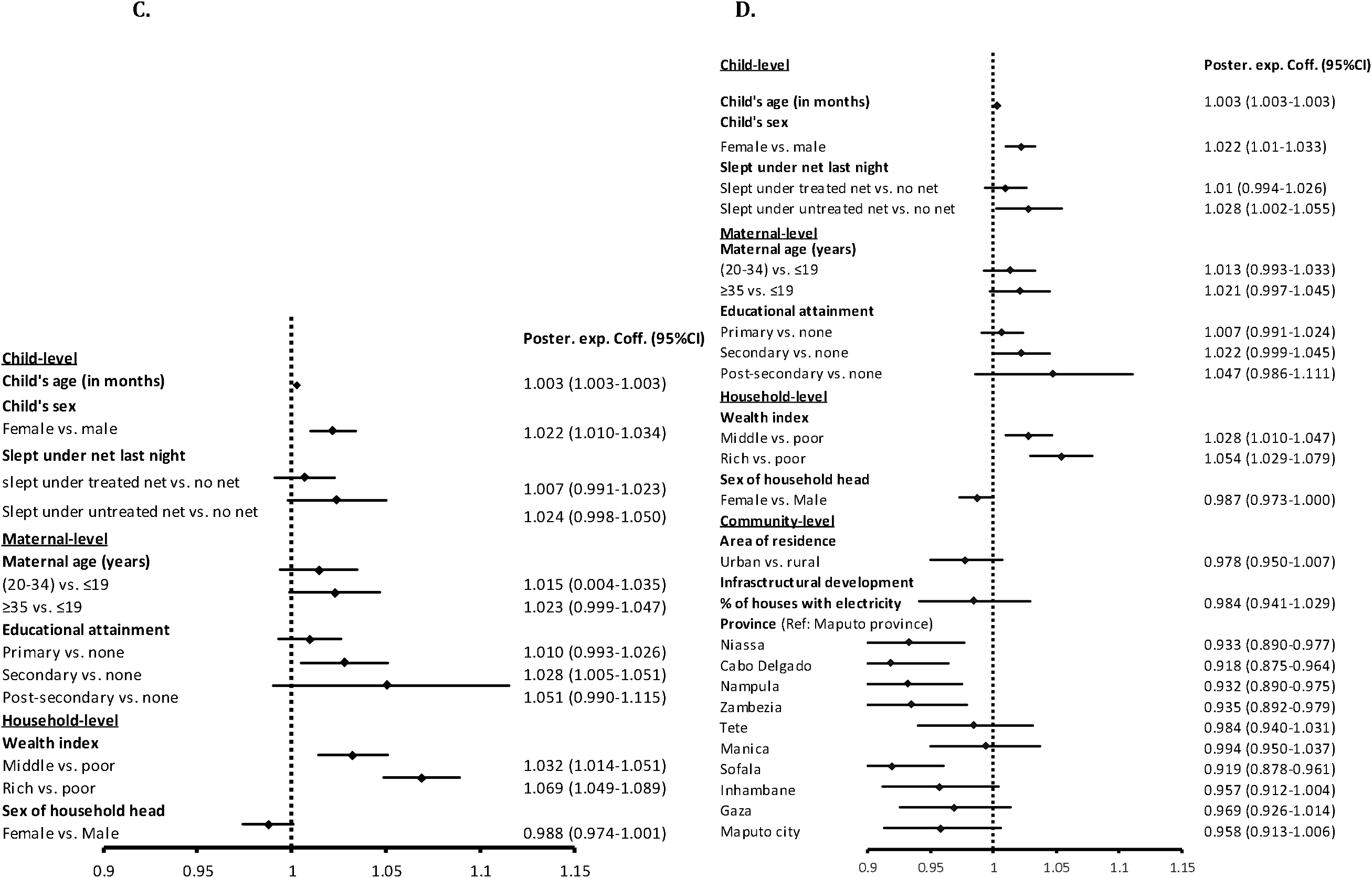
Parameter estimates (fixed-effects) of the association between childhood haemoglobin concentration and child, maternal, household, and community-level factors A. Model 1 (child-level factors) B. Model 2 (child and maternal level factors) C. Model 3 (child, maternal and household level factors) D. Model 4 (child, maternal, household, and community-level factors), Mozambique MIS, 2018

With an average DIC of −2490 for model 3 and model 4, adding household and community-level factors improved the models (Table 2). Although, both models had similar average DIC value, model 4 was selected because of the further reduction in variance across the communities, with the addition of community-level factors.

**Table 2:** Random-effect, efficiency and Gelman-Rubin convergence diagnostic summaries

| Variables | Model 0<br>(Empty) | Model 1<br>(child) | Model 2<br>(child and<br>maternal) | Model 3<br>(child, maternal<br>and household) | Model 4<br>(child, maternal, household<br>and community) |
| --- | --- | --- | --- | --- | --- |
| **Variance (log)** |  |  |  |  |  |
| Community | 0.0043 | 0.0041 | 0.0036 | 0.003 | 0.0025 |
| Household | 0.003 | 0.0059 | 0.0056 | 0.0033 | 0.003 |
| Maternal | 0.0026 | 0.004 | 0.0042 | 0.003 | 0.0034 |
| **MOR** |  |  |  |  |  |
| Community | 1.0643 | 1.0627 | 1.0587 | 1.0534 | 1.0487 |
| Household | 1.0534 | 1.0757 | 1.0737 | 1.0561 | 1.0534 |
| Maternal | 1.0496 | 1.0619 | 1.0635 | 1.0534 | 1.0570 |
| **ICC** |  |  |  |  |  |
| Community (%) | 0.13 | 1.2 | 0.11 | 0.09 | 0.08 |
| Household (%) | 0.22 | 0.3 | 0.28 | 0.19 | 0.17 |
| Maternal (%) | 0.3 | 0.42 | 0.41 | 0.28 | 0.27 |
| **PCV** |  |  |  |  |  |
| Community (%) | Ref. | 4.65 | 16.28 | 30.23 | 41.86 |
| Household (%) | Ref. | -96.6 | -86.6 | -10 | 0 |
| Maternal (%) | Ref. | -53.85 | -61.54 | -15.39 | -30.77 |
| Average efficiency (%) | 22.84 | 10.66 | 13.03 | 16.47 | 13.55 |
| Average acceptance rate (%) | 69.76 | 69.52 | 69.99 | 69.29 | 69.67 |
| Maximum Gelman-Rubin Rc | 1.10 | 1.08 | 1.02 | 1.02 | 1.09 |
| Average DIC | -2100 | -782.24 | -784.98 | -2490 | -2490 |

MOR: Median odds ratio, ICC: Intraclass correlation, PCV: Proportional change in variance, DIC: Deviance Information Criteria

**Table 2:** Areas with significant clustering of children with low haemoglobin concentration after adjusting for child, maternal, household and community-level factors, Mozambique MIS, 2018

| Province /Administrative boundaries | District |
| --- | --- |
| Niassa-Cabo Delgado-Nampula | Nipepe, Lalaua, Balama, Namuno |
| Gaza-Inhambane | Mabote, Funhalouro, Chigubo |
| Zambezia-Nampula | Malema, Ribaue, Alto Molocue, Gurue |
| Manica | Vanduzi |
| Inhambane | Morrumbene, Maxixe |

### Measures of variance and clustering in multilevel Bayesian regression

For the intercept-only model (Model 0), Table 2 shows low intra-cluster correlations (ICC), implying an existence of weak maternal (0.3%), household (0.2%) and community effects (1.3%) (i.e., small variations in childhood anaemia between women, households and communities). Sequential inclusion of child, maternal, household and community-level factors further reduced the variability (ICC) in childhood anaemia—i.e., 0.27%, 0.17% and 0.08% of the variances were unexplained at the maternal, household and community levels, respectively. In the final model, close to half of the variance across the communities were explained by the child-, maternal-, household-, and community-level factors (PCV=41.86%). Importantly, as determined by proportional change in variance, 37.21% of the community-level variance of childhood anaemia were accounted for by the joint effects of maternal-, household-, and community-level factors. In the same vein, the unexplained heterogeneity between the communities decreased from MOR of 1.064 in the empty model to 1.049 in the final model (Model 4), hence there was little residual variation in childhood anaemia across the communities. Thus, if a child moves to a community with a higher probability of being anaemic, the median risk of having anaemia increases by 1.1 times.

### Spatial variation of childhood anaemia

After accounting for uncertainty and variance instability with spatial empirical Bayesian smoothing map, the smoothed prevalence of childhood anaemia by province ranged from 0.72 (Maputo city) to 0.84 (Cabo Delgado, Zambezia, Niassa and Nampula), with a national smoothed mean (SD) of 0.78 (±0.05) (Figure 2a). Children living in four provinces had excess risk of anaemia relative to the national average: 1.07 in Cabo Delgado, 1.05 in Nampula, 1.04 in Zambezia and 1.02 in Sofala (Figure 2b).

**Figure 2:**
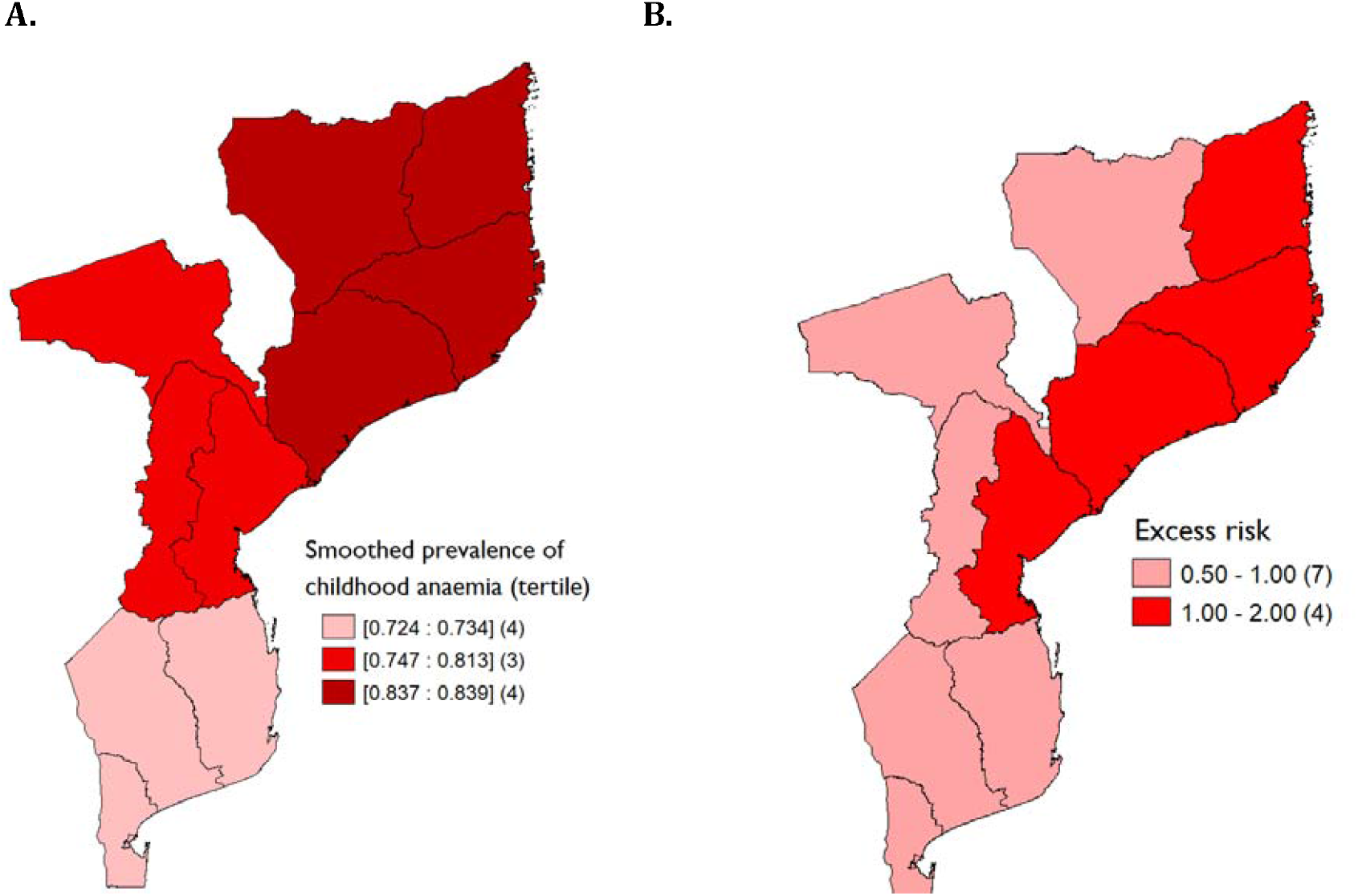
Spatial distribution of childhood anaemia by province in Mozambique A. Spatial empirical Bayesian smoothed prevalence B. Excess risk, Mozambique MIS, 2018

There was significant clustering of prevalence of childhood anaemia across the communities. Strong spatial clustering of prevalence of childhood anaemia was observed at the provincial level (Moran’s I=0.64, p-value=0.005), while clustering was weaker at the community-level (I=0.1, p-value=0.001) (Supplementary Figure S6). Forty-four communities (20%) clustered to form cold spots of childhood anaemia i.e., clustering of communities with relatively lower prevalence of childhood anaemia compared to average of spatial units. All the cold spots were in the southern part of the country—13 communities in Maputo province, 12 in Gaza, 11 in Maputo city, and 8 in Inhambane. Conversely, all the 47 (21.4%) communities that formed the hot spots with relatively higher prevalence of childhood anaemia (compared to the community average) were found in northern Mozambique—16 communities in Cabo Delgado, 15 in Nampula, 10 in Zambezia and 6 in Niassa. Also, there were 18 communities (8.2%) in the low-high clusters (communities with low prevalence of childhood anaemia adjacent to hot spots)—located in Nampula and Zambezia. Twenty-seven communities (12.3%) recorded higher prevalence and were adjacent to communities with low prevalence (high-low clusters).

Figure 3 shows the crude and adjusted predictive maps of community-level random effects of posterior mean haemoglobin concentration from the multilevel Bayesian regression. Compared to the crude predictive map (Model 0), the adjusted map (Model 4) shows markedly reduced childhood anaemia and some residual spatial variation across the communities after controlling for child, maternal, household and community-level factors, but disparities persist along the administrative boundaries. Most communities around Gaza-Inhumbane boundary, Niassa-Cabo Delgado-Nampula tripoint and Zambezia-Nampula boundary, and in Manica and Inhambane provinces have negative posterior means, indicating increased probability for childhood anaemia. Table 2 shows the specific districts with significant clustering of children with low haemoglobin concentration after adjusting for the independent variables.

**Figure 3:**
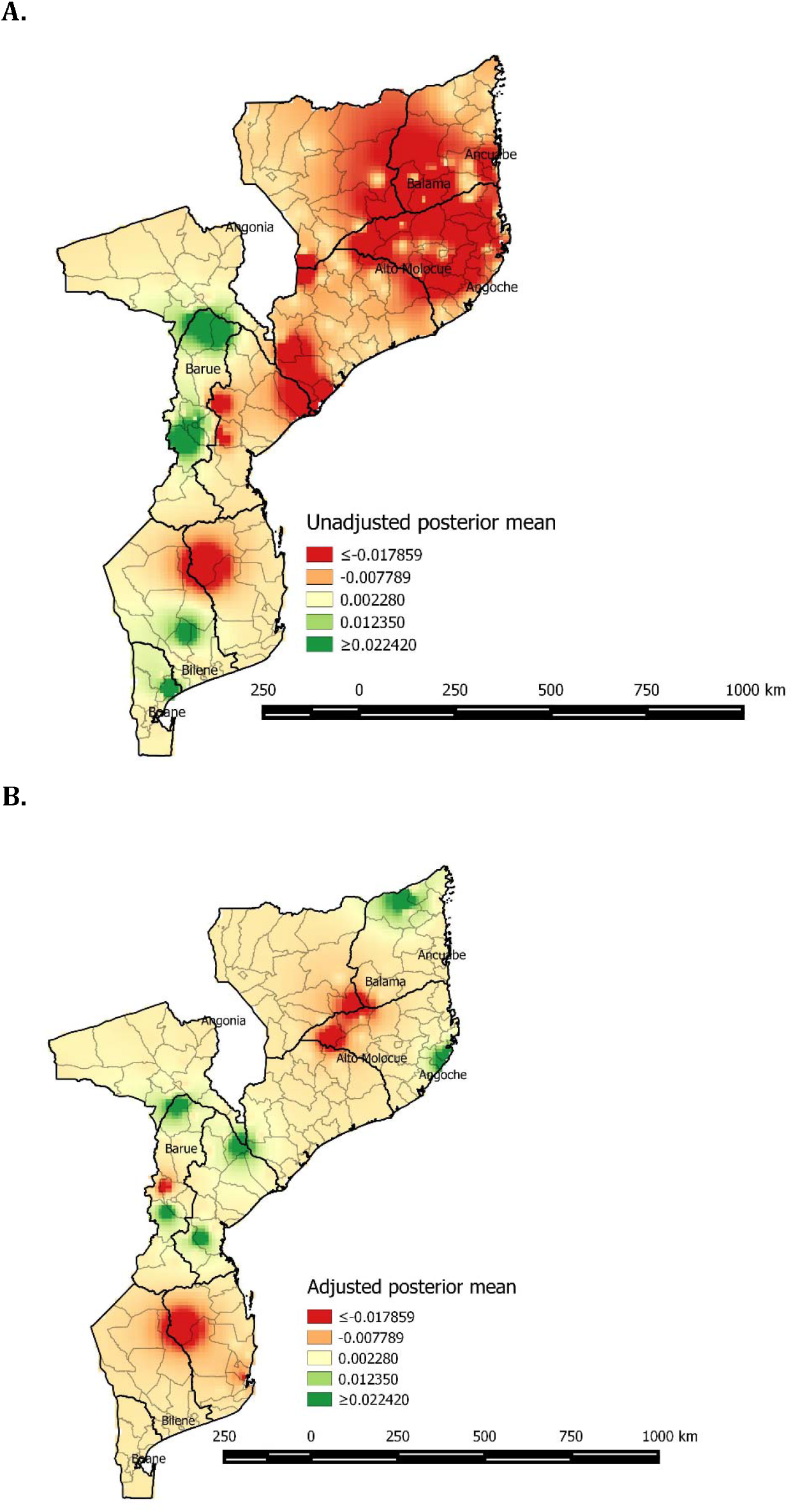
Spatial distribution of posterior means of haemoglobin concentration across communities in Mozambique A. Crude map B. Adjusted map, Mozambique MIS, 2018 Color code: Red denotes high risk, green denotes lower risk

## Discussion

This study examined the influence of individual-(child and maternal), household-, and community-level factors on the geographical distribution of anaemia among children aged 6-59 months in Mozambique using the 2018 Malaria Indicator Survey. A key finding is that childhood anaemia is highly prevalent although showed spatial variation across the communities and provinces in Mozambique and were related to socioeconomic factors. Within the study population, Children with anaemia tended to be younger, males, and at risk of having malaria because they were not sleeping under mosquito nets. Furthermore, there is evidence that children from poor families, those living in female-headed households were at greater risk of developing anaemia, and several provinces (located majorly in northern Mozambique)—Cabo Delgado, Nampula, Niassa, Sofala and Zambezia were prone to anaemia.

Our analysis showed an overall national childhood anaemia prevalence of 78-80.3%, mostly moderate anaemia at the time of survey. With such a high prevalence of anaemia in children— exceeding 40% across the provinces—childhood anaemia remains a severe public health threat in Mozambique, as indicated by the population benchmark by WHO for classifying prevalence of anaemia as a problem of public health significance.^(26)^ Although, estimated national prevalence in this report is higher than the 69% previously reported,^(16)^ the observed pattern of variation is consistent with the 2011 Mozambique DHS report— Maputo province with the lowest prevalence and districts within Cabo Delgado, Zambezia and Nampula the highest.^(16)^ It is also important to note that, with the exception of Maputo province, which had prevalence of severe anaemia of 0.28%, other provinces had exceeded the recommended 2% (ranged from 2.88% in Maputo city to 12.17% in Cabo Delgado). As recommended by the International Nutritional Anaemia Consultative Group (INACG),^(37)^ the policy implication of this finding is that, training and supervision of healthcare workers in the primary care centres on early detection and treatment/referral of children with severe anaemia should be intensified.

After addressing potential confounders, we observed higher risk of childhood anaemia in communities located near administrative borders. Most of the communities were found around the tri-provincial border of Niassa-Cabo Delgado-Nampula (i.e., Nipepe, Lalaua, Balama, Namuno); district boundaries of Mabote-Funhalouro-Chigubo in Gaza and Inhumbane provinces; Malema, Ribaue, Alto Molocue, Gurue in Zambezia and Nampula provinces; Malema, Ribaue, Alto Molocue, Gurue in Manica province and Morrumbene, Maxixe in Inhambane province. This variation was mainly due to maternal-level factors (maternal age and educational attainment), household-level factors (gender of household head and household wealth) and community-level factors (place of residence and infrastructural development). Child-related factors were observed to have lesser effect on the geographical distribution of childhood anaemia in Mozambique. This study reaffirms the need to prioritize community health, especially maternal and child health, around district, provincial and international boundaries. The strong link between international and provincial borders, and health outcomes have been documented in previous studies for maternal malnutrition,^(38)^ onchocerciasis,^(39)^ and cholera outbreak.^(40)^

Several studies have linked childhood anaemia with limited household resources because household wealth is a major determinant of food (in)security and health care utilization.^(41–43)^ Our results suggest that childhood anaemia decreases with increasing household wealth. As expected, childhood anaemia was observed to be higher in provinces with higher percentage of households living in poverty. Furthermore, our analysis found that anaemia risk reduces as child grows older. This finding is comparable to similar studies elsewhere.^(41,42,44)^ Tsai et al.,^(45)^ and Paoletti et al.,^(46)^ raised concerns about low iron concentration in human breast milk and in most traditional diets, which might be insufficient to meet the daily micro-nutrient requirements for rapid growth during the period of weaning (after 6 months). This is partly indicative of the inability of households to introduce iron-rich complementary foods after 6 months, arising from their low socioeconomic status.

A potential intervention to reduce the prevalence of anaemia among children is effective malaria prevention. As a key malaria intervention, we found evidence that mosquito bed net use lowers the risk of anaemia. Experiences in Ghana have indicated that anaemia accounted for more than half of malaria-related deaths.^(47)^

One of the strengths of our study is the investigation of determinants conceptualized and applied at four-levels: community, household, maternal, and child. To the best of our knowledge, no published literature has taken into consideration the contextual effects at the community-, household-, and maternal-levels. Also, we used Bayesian methods to simulate the nationally representative dataset from the latest MIS for Mozambique. With Bayesian methods, uncertainty around point estimates are minimized, hence generating more accurate models. Using multilevel regression and spatial analysis, our study provides new findings about the geographical distribution (i.e., vulnerable groups at the borders) and social determinants of anaemia among children aged 6-59 months in Mozambique. As this study provides analysis in a more granular level (community and district), it provides baseline analysis that can guide general and targeted implementation of the 2025 Mozambique National Development Plan for optimal use of resources. A limitation of this study is that the cross-sectional nature limits causal inferences. Also, we could not directly estimate the effects of recent malaria illness, food insecurity, HIV and intestinal worm infestation because they were not available in the dataset. To overcome the challenge of missing variables, we used mosquito bed net usage (as a proxy for malaria illness), household sanitation (as a proxy for worm infestation) and household wealth index (as a proxy for food insecurity).

This study provides evidence that anaemia among children aged 6-59 months is a severe public health threat across the provinces in Mozambique. It also identifies inequity in childhood anaemia—worse among communities living close to the provincial borders. We recommend interventions that would generate income for households, increase community-support for households headed by women, improve malaria control, build capacity of healthcare workers to manage severely anaemic children and health education for mothers. More importantly, there is need to foster collaborations between communities, districts and provinces to strengthen maternal and child health programmes for the severely affected areas.

### Postscript

Given this paper is authored by four health professionals representing two continents, three countries and a wide range of experiences, we wish to declare the standpoint from which we write (“pose”) and the audience that we intend to reach (“gaze”). The standpoint taken is primarily African, Asian, and trained in both African and North American education and clinical training systems. We bring both a local (Mozambican, African) and foreign perspective. The foreign perspective (N.M.) however is tempered by many years of working with local colleagues. Our gaze is ‘local’ in that we are writing for the audience of policy-makers, health care professionals, government leaders in Mozambique. For a fuller explanation of ‘pose’ and ‘gaze’ intersected by local and foreign actors producing research reports such as the present, please see: Abimbola S. The foreign gaze: authorship in academic global health. BMJ Global Health 2019;4:e002068. doi:10.1136/bmjgh-2019-002068.

## Data Availability

The dataset for this study is publicly available at the DHS website.

https://dhsprogram.com/data/Dataset-Availability-Status-Overview.cfm

## Acknowledgements

The authors wish to thank DHS Programme for granting access to the dataset. We also acknowledge the translation support provided by Ms. Emiliana Bomfim.

## Data availability and funding

The dataset for this study is publicly available at the DHS website. D.A. is funded by Mozambique-Canada Maternal Health Project.

## Competing interest

The authors declare no conflict of interest or stand to benefit from this work in any material manner.

## Authors’ contributions

Please see the Postscript as well. Authors contributed to the study in the following manner: conceptualization, N.M., D.A.; methodology, N.M., D.A.; formal analysis, D.A.; interpretation and writing—review, editing, N.M., D.A., M.M., S.C.; supervision and resources, N.M. All authors have read and agreed to the submission of the manuscript.

## Notes

### Competing Interest Statement

The authors have declared no competing interest.

### Clinical Trial

Not a clinical trial.

### Funding Statement

Global Affairs Canada, D-002085/P001061

### Author Declarations

This is a secondary analysis of data available online where the datasets are de-identified of the respondents' personal information, hence no ethical approval was required for this specific study. However, prior to the commencement of the primary survey, ethical clearances were obtained by the Demographic and Health Survey (DHS) team from National Committee for Bioethics in Health of Mozambique. Also, written informed consent was obtained from all mothers during the field work. Following registration, administrative access to the dataset was granted by the DHS team, United States. The survey data files were provided at no cost for academic research.

